# Psychological consequences of the Flint Water Crisis: A systematic review

**DOI:** 10.1101/2020.05.30.20117952

**Authors:** Samantha K. Brooks, Sonny S. Patel

## Abstract

Given the COVID-19 outbreak, these unprecedented times pose many urgent dilemmas about how to support the mental health of communities. The goal of this research is to summarize existing literature on the mental health impact of the recent Flint Water Crisis. In March 2020, we searched five databases for literature exploring the psychological consequences of the crisis. Main findings were extracted. 132 citations were screened and eleven included in the review. Results suggest a negative psychological effect caused by the water crisis, including anxiety and health worries, exacerbated by lowered trust in public health officials, uncertainty about the long-term impacts of the crisis, financial hardships, stigma, and difficulties seeking help. There was evidence that concerns about tap water continued even after the state of emergency was lifted. With a possible compounded effect to residents of Flint with the recent COVID-19 pandemic, the results highlight the need for more resources for psychological health interventions in Flint as well as a need for local governments and health authorities to regain the trust of those affected by the Flint Water Crisis.

## Introduction

The city of Flint is the urban centre of Genesee County, Michigan, USA with a population of over 95,000 according to 2018 estimates (United States Census Bureau, 2019). On 25th April 2014, Flint changed its municipal water supply source from Lake Huron to the Flint River as a cost-saving measure (Adams, 2014). However, the Flint River water was not treated with corrosion control chemicals to ensure the more acidic river water did not cause corrosion of water distribution pipes. By summer 2014, Flint residents had begun reporting changes in the smell, taste and appearance of their water as well as health effects such as skin rashes and hair loss. However, officials insisted the water was safe and dismissed the idea of a link between water quality and health problems, and residents continued to use tap water. Meanwhile, the supply pipes continued corroding, leaching lead into the water (Kennedy, 2016).

In September 2015, water experts discovered very high levels of lead in the tap water of some Flint homes (Flint Water Study, 2015), and a local paediatrician found increases in children’s blood lead levels corresponding with the time of the switch in water sources (Hanna-Attisha et al., 2016). Finally, the state re-evaluated its water-testing data, discovering elevated levels of contaminants including bacteria and lead in Flint’s drinking water, and concluded that the water was, in fact, unsafe (Kennedy, 2016). Although the water source was switched back to the Lake Huron source in October 2015, a state of emergency was declared at both the state and federal level in January 2016 which was in effect until August 2016. Despite the state of emergency having been lifted, according to media reports many residents remain fearful of Flint’s water, feel they have not received any explanation for why the crisis was allowed to happen and still lack trust in public health officials (Renwick, 2019).

Naturally, the crisis has raised concerns about the physical health of Flint residents. Lead exposure can lead to high blood pressure, heart disease, damage to the brain and kidneys, and infertility (Agency for Toxic Substances and Disease Registry, 2007). Lead exposure is particularly harmful for children, putting them at greater risk of brain and nervous system damage, slowed development and behavioural problems (Grandjean & Landrigan, 2014). In addition to physical health concerns, there are also potential mental health consequences of the crisis which cannot be overlooked. Previous research suggests that experiencing a disaster or public health emergency – particularly one that is human-induced – can lead to mental health disorders and substantial negative effects on levels of stress (Makwana, 2019). Psychological consequences can occur not only during and in the short-term aftermath of a disaster or crisis but can also affect both adults and children for years after (Bland et al., 1996; Bolton et al., 2000).

Residents of Flint may be particularly at risk of adverse mental health consequences due to the city’s long-standing social and economic vulnerabilities: almost half of Flint’s residents live below the federal poverty level (United States Census Bureau, 2019) and Flint has consistently been rated one of the most violent cities in the US (Harris, 2010; Warner et al., 2013). Flint also has a long history of racial segregation, with environmental racism believed to be a contributor to the water crisis (Michigan Civil Rights Commission, 2017). Disadvantaged communities are more likely to be vulnerable to adverse mental health outcomes after a disaster or emergency and have more barriers to treatment (SAMHSA, 2017) so the mental health of Flint residents is of particular concern.

Despite the risk to Flint residents in terms of their mental health, the psychological impact of the crisis has received little attention in the literature, and human-focused recovery efforts have been minimal in comparison to the recovery of physical infrastructure (Kruger et al., 2017a). But the mental health impact from the Flint Water Crisis may be a long-lasting legacy for generations to come in the community, as has been seen in other post-disaster communities: for example, the fallout of the Chernobyl Nuclear Power Plant in Pripyat, Ukraine led to gaps in providing mental health care and research to disaster-affected communities since 1986, and this mental health impact has been labelled as the largest public health problem caused by the accident (The Chernobyl Forum, 2006). In fact, the media have frequently compared the Flint situation with the Chernobyl disaster (Al-Sibai, 2019; McFarland, 2019), while researchers have labelled the Flint Water Crisis more ‘insidious’ than Chernobyl (Campbell et al., 2016), which causes concern as to the potential long-lasting impact of the crisis. Past disasters in other communities have shown the importance of resilience, which includes the ability to return to self-sufficiency and sustain relatively stable psychological and physical functioning after a traumatic event (Abir et al., 2016), which highlights the importance of considering the level of resilience in Flint’s community and how this can be improved. It is important that the psychological consequences of the crisis are not overlooked – particularly now, with the ongoing coronavirus (COVID-19) pandemic. Flint has the highest number of COVID-19 cases in Genesee County: as of 04 May 2020, Flint had recorded 644 cases (representing 39.5% of Genesee County cases), followed by Flint Township with 125 (7.6% of Genesee County’s cases) (Genesee County Health Department, 2020). The psychological consequences of the unprecedented lockdown of communities in order to reduce transmission is likely to be substantial (Brooks et al., 2020) and may be particularly so for communities which have yet to fully recover from a past crisis and are now faced with another one. Taking account of the lessons learned from the fallout from the Chernobyl disaster and with the current situation with the COVID-19 pandemic, applying best practices are important for responding and serving the affected populations.

This paper aimed to systematically review the published literature on the psychological impact of the Flint water crisis, specifically focusing on i) what is the mental health impact of the crisis? and ii) what are the factors associated with this impact?

## Method

### Search strategy

On 27^th^ March 2020, the following search strategy was used to search titles and abstracts in five databases (Medline, PsycInfo, Embase, Global Health and Web of Science) from inception to 2020 Week 12:

1. Flint
2. Water
3. Mental health
4. Behavioural health
5. Behavioral health
6. Psycholog*
7. Consequence*
8. Impact*
9. 3 OR 4 OR 5 OR 6 OR 7 OR 8
10. 1 AND 2 AND 9

### Inclusion criteria

To be included in the review, studies had to: i) include primary data; ii) be published in peer-reviewed journals; iii) be written in English; and iv) report on either the psychological consequences of the Flint water crisis or factors associated with psychological outcomes as a result of the crisis.

### Screening

One author (SKB) ran the search strategy on all databases and downloaded resulting citations to EndNote version X9 (Thomson Reuters, New York, USA) where duplicates were automatically removed. The same author then screened all titles for relevance to the review, excluding any which were clearly not relevant. This was followed by screening of abstracts. The full texts of all citations still remaining were obtained and screened for relevance against the inclusion criteria. Finally, the reference lists of included papers were hand-searched for additional relevant studies.

### Data extraction and synthesis

A spreadsheet was designed in order to systematically extract data from the literature. The following data was extracted: year of study, design and measures used, number of participants, demographic information of participants, and key results. Data extraction was carried out by one author (SKB). Thematic analysis (Braun & Clarke, 2006) was used to synthesise the data by coding it and organising it into themes.

## Results

Database searches yielded 216 articles. After 84 duplicates were removed, 132 articles remained for screening. Title screening removed 101 of these; abstract screening removed another 15; and 16 full texts were reviewed. Five were excluded, leaving eleven included in the review (Figure I). Table I summarises the design and participant information for each study and Table II summarises the evidence for each of the themes found in the literature.

**Table I.** Characteristics of included studies

| **Study** | **Design and measures** | **Participants** |
| --- | --- | --- |
| Cuthbertson et al. (2016) | 12 monthly surveys with questions about changes in 30 behavioural health related issues and open-response questions about new issues and community events since the previous survey. | 30 expert panellists from health services, substance use prevention, health-related NGOs, disability service organisations, schools and researchers in Genesee County. |
| Fortenberry et al. (2018) | Cross-sectional; 2-page | 180 Flint residents over the |
|  | questionnaire addressing household demographics, communications, water sources and uses, behavioural health concerns, access and perceived barriers to behavioural health services, chronic disease diagnoses, self-reported physical health consequences, and individual-level behavioral health concerns. Questions from PHQ-2 and GAD-2 to assess depression and anxiety symptoms. | age of 18. Of households interviewed: 88.3% had one or more members aged 21-64, 60.5% had one or more 20 or younger, 25.3% had one or more 65+. Average number of people living in household was 3 (range 1-10). Mean age 49.5 years. 69.2% female. |
| Gray et al. (2017) | 2-hour modelling workshops using Mental Modeler software and follow-up cultural consensus survey. | 42 in workshops; 137 survey participants. |
| Heard-Garris et al. (2017) | Cross-sectional; survey focusing on exposure, consequences and coping strategies. | 133 drug users; mean age 26.1; 52% female. |
| Kruger et al. (2017a) | Cross-sectional; Short Screening Scale for PTSD | 786 Genesee County residents; mean age 51; 72% female. |
| Kruger et al. (2017b) | Cross-sectional; survey including mental and physical health items from the CDC's Behavioral Risk Factor Surveillance System with items on self-reported health and mental health. | 277 residents of Flint aged 18+; 69% female; mean age 48 (range 18-94). |
| Kruger et al. (2017c) | Cross-sectional; Survey asking about sleep quality | 834 Genesee County residents. |
|  | overall during the past month and sleep length during a typical night. |  |
| Kruger et al. (2018) | Cross-sectional; Survey including several scales relevant to life history theory. | 701; mean age 49, range 18-94; 70.9% female. |
| Morckel & Terzano (2019) | Cross-sectional; Survey asking about levels of concern about water crisis, likelihood of leaving the city, perception of water quality, concerns about housing, levels or trust in government, and basic health indicators. | 405 Flint residents. |
| Singer et al. (2017) | Community-based 'mental modelling' workshops using Fuzzy Cognitive Mapping and Mental Modeler software to capture beliefs about causes, consequences and solutions to the water crisis; survey asking participants to rank the trustworthiness and usefulness of information sources. | 36 Flint residents and officials. |
| Sneed et al. (2020) | Community Assessment for Public Health Emergency Response (CASPER) assessments over three years; Survey to assess household and individual level self-reported behavioural health concerns, access to behavioural health services, physical health concerns, and | 180 in 2016, 176 in 2017, 193 in 2018. |
|  | water-related resource needs. |  |

**Table II.**
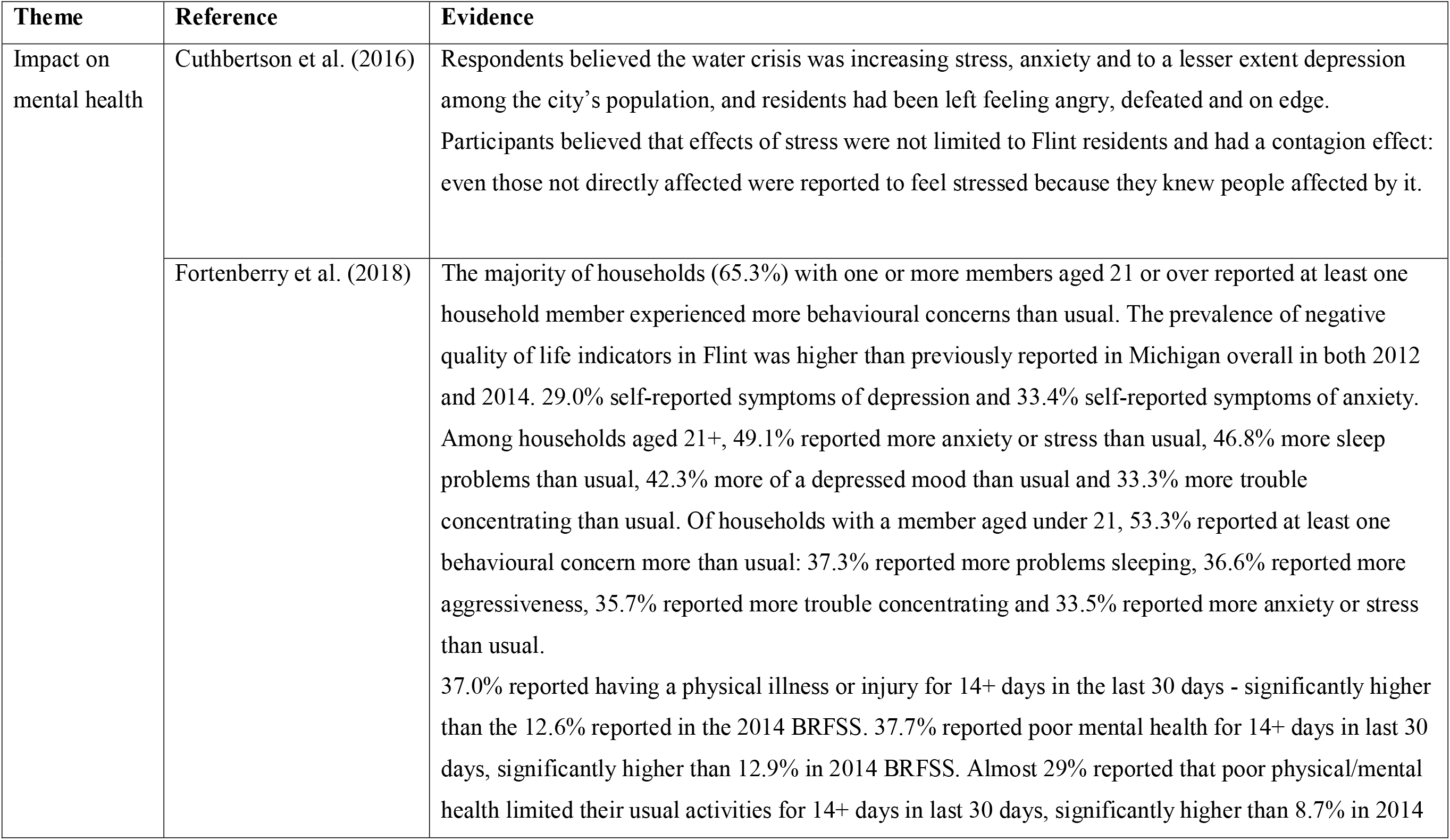
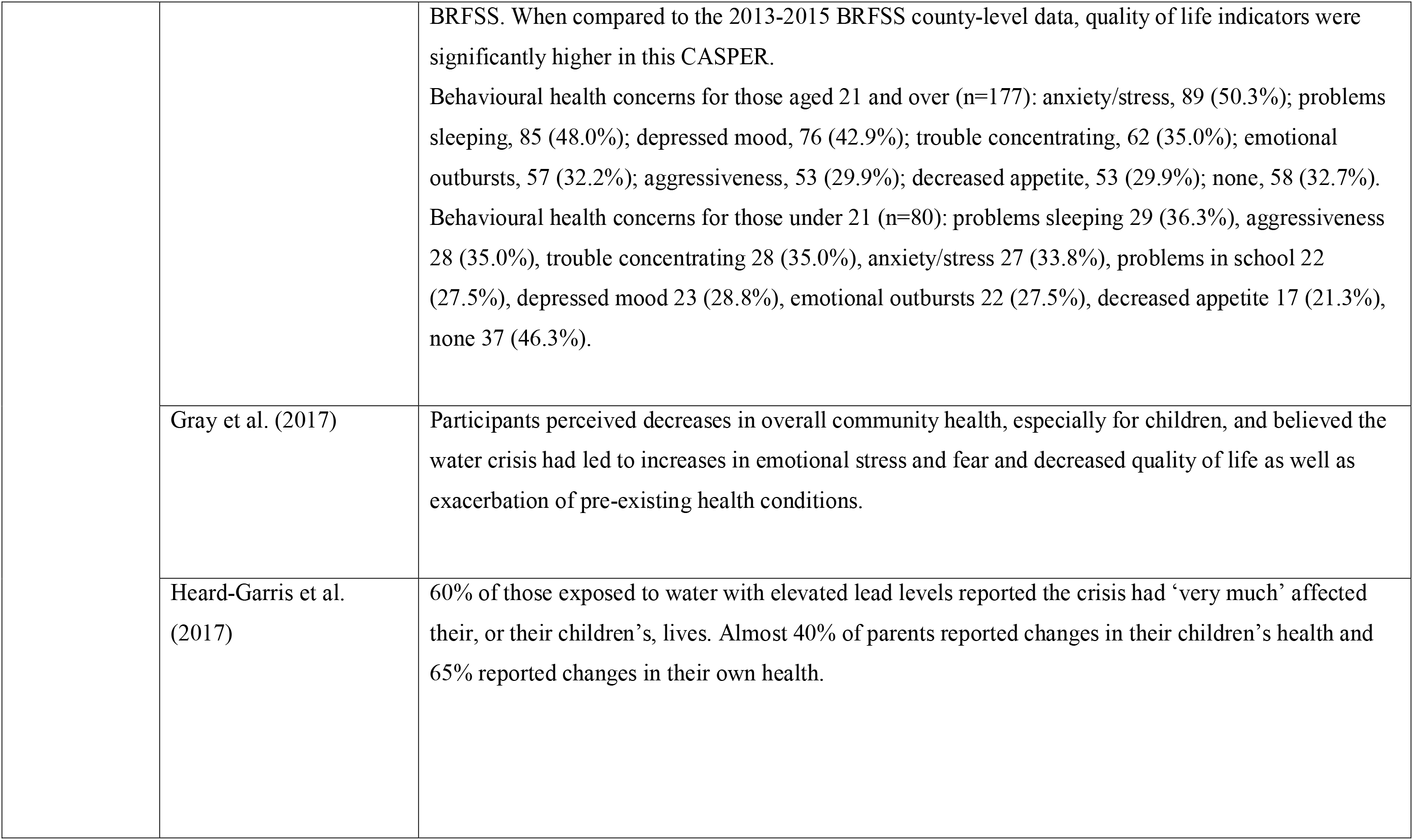
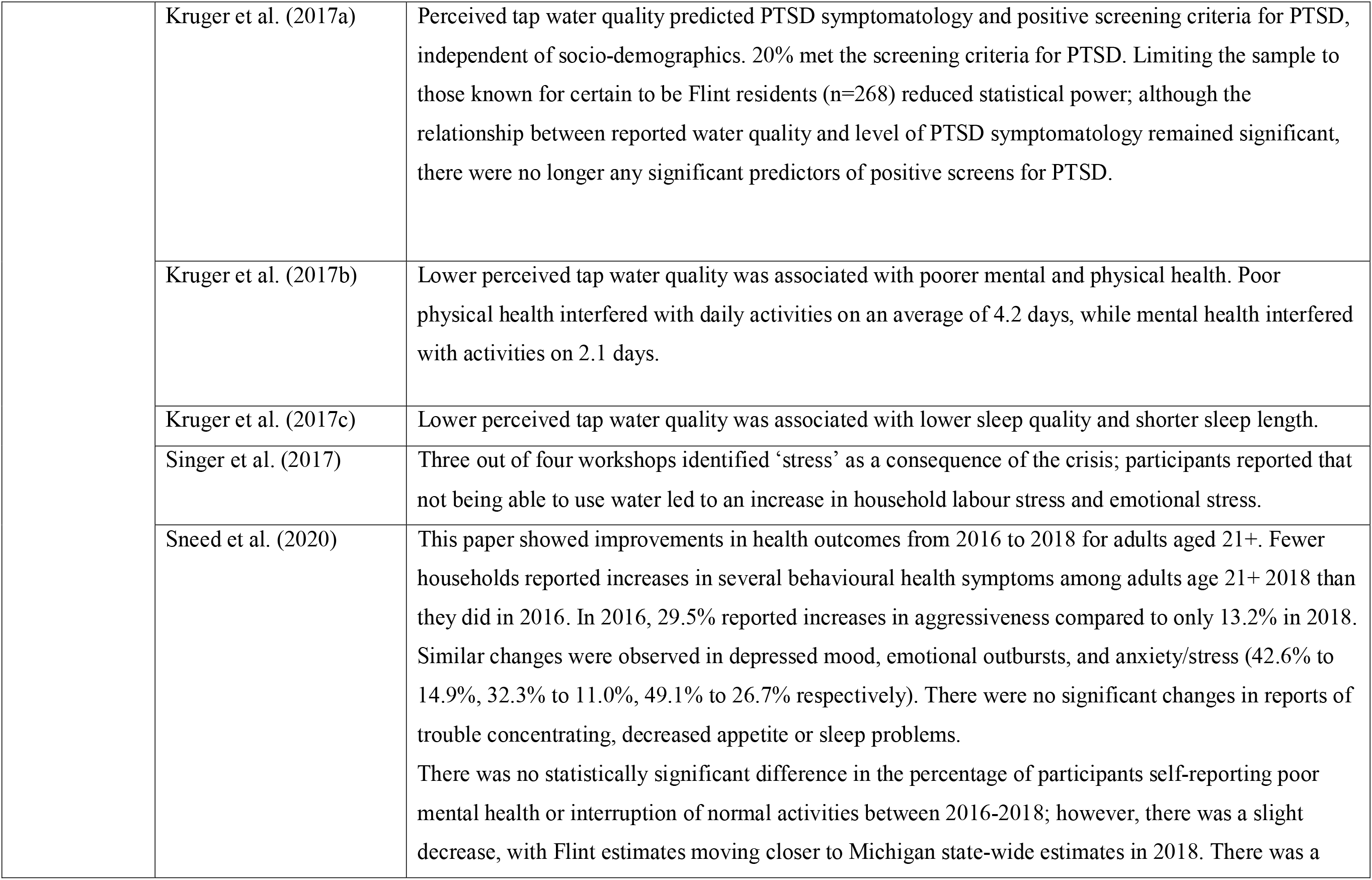
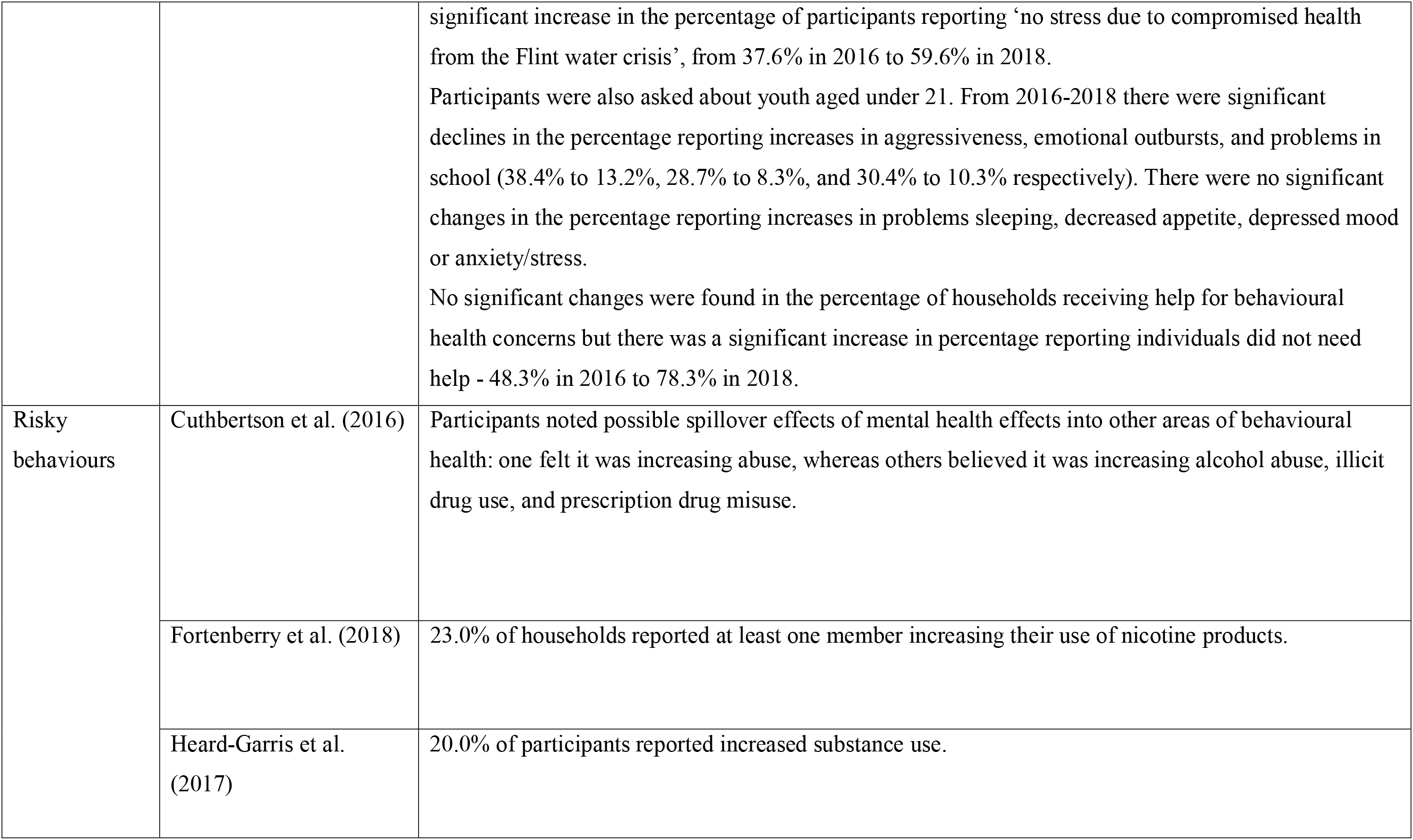
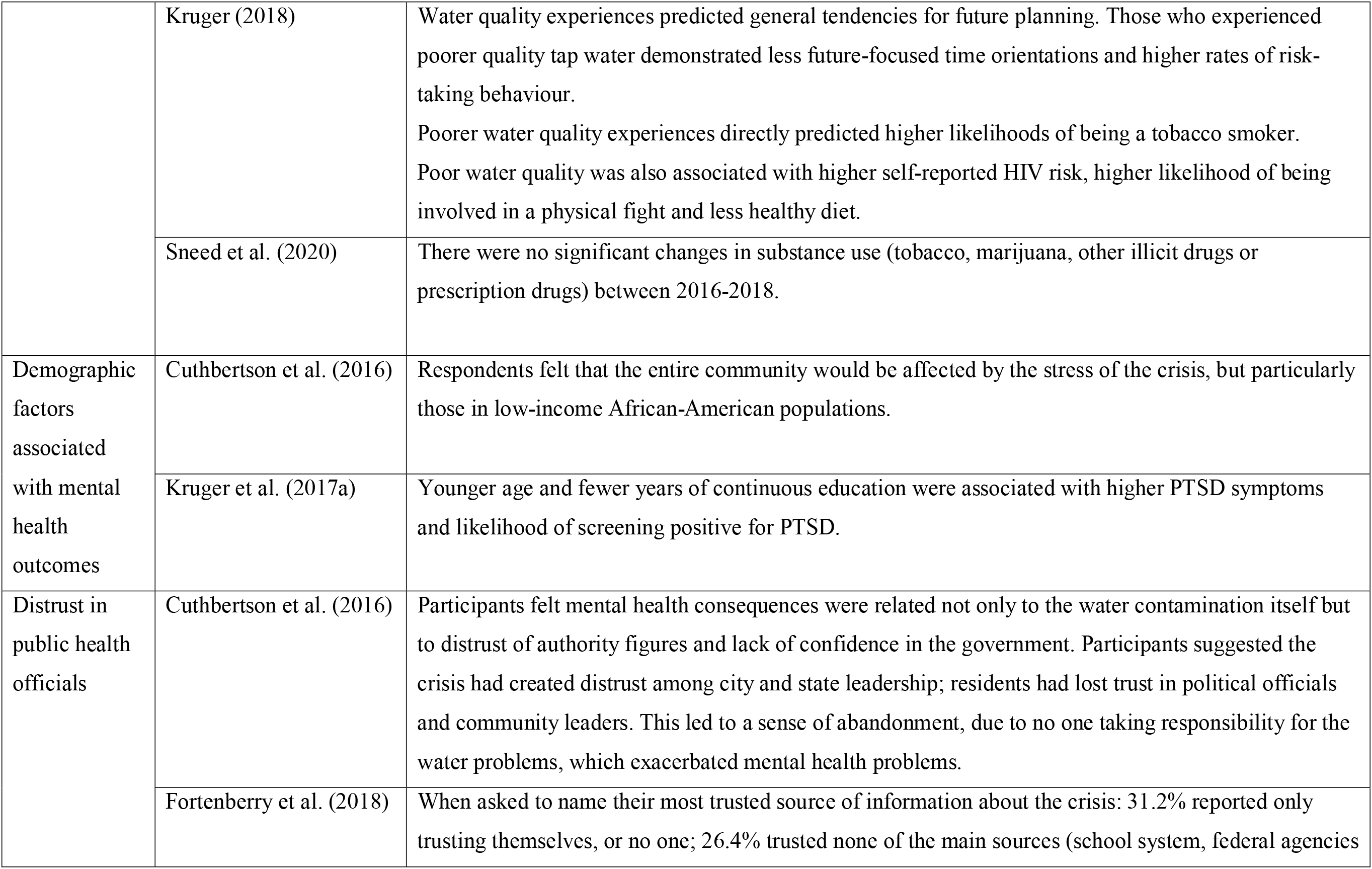
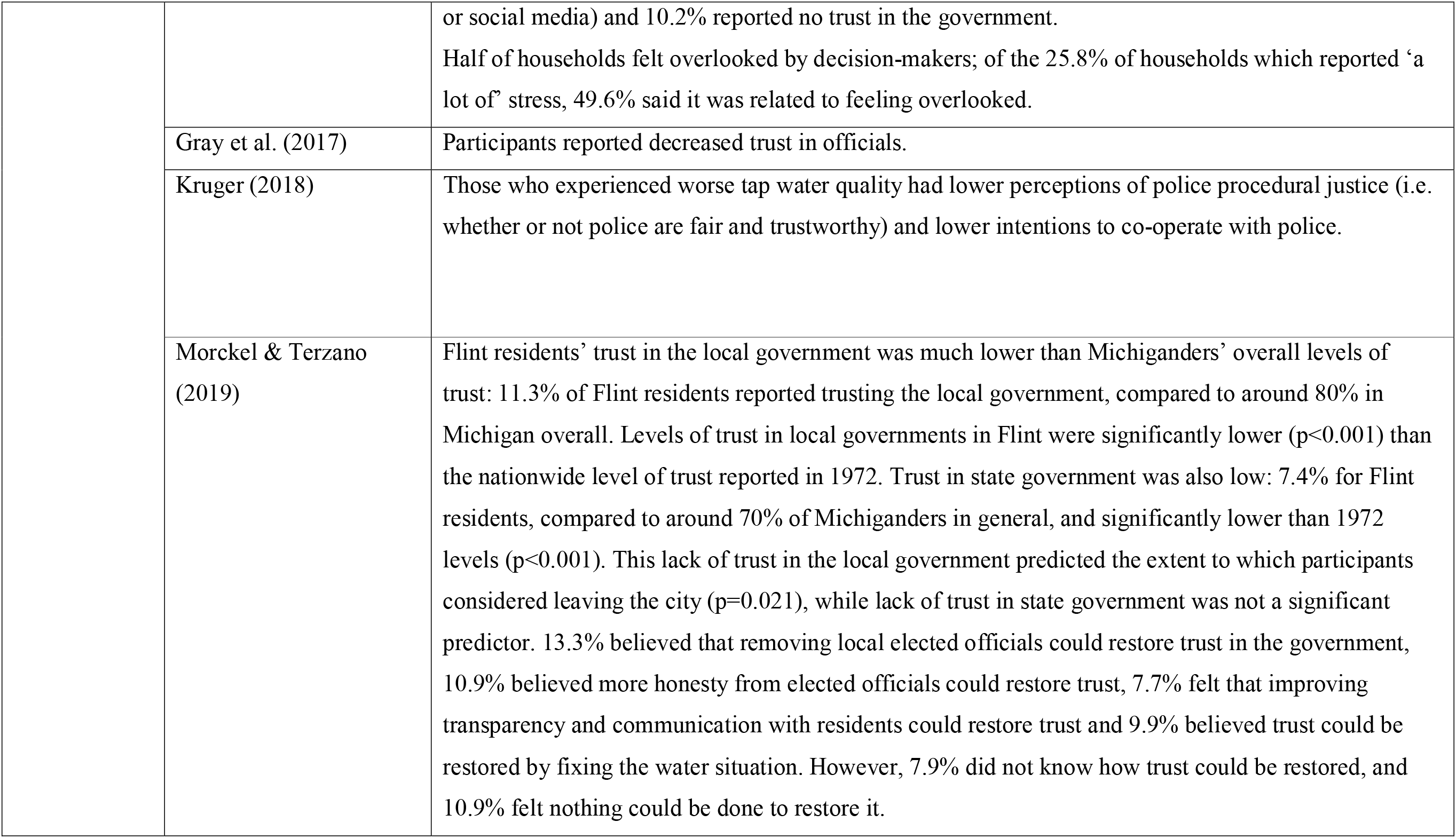
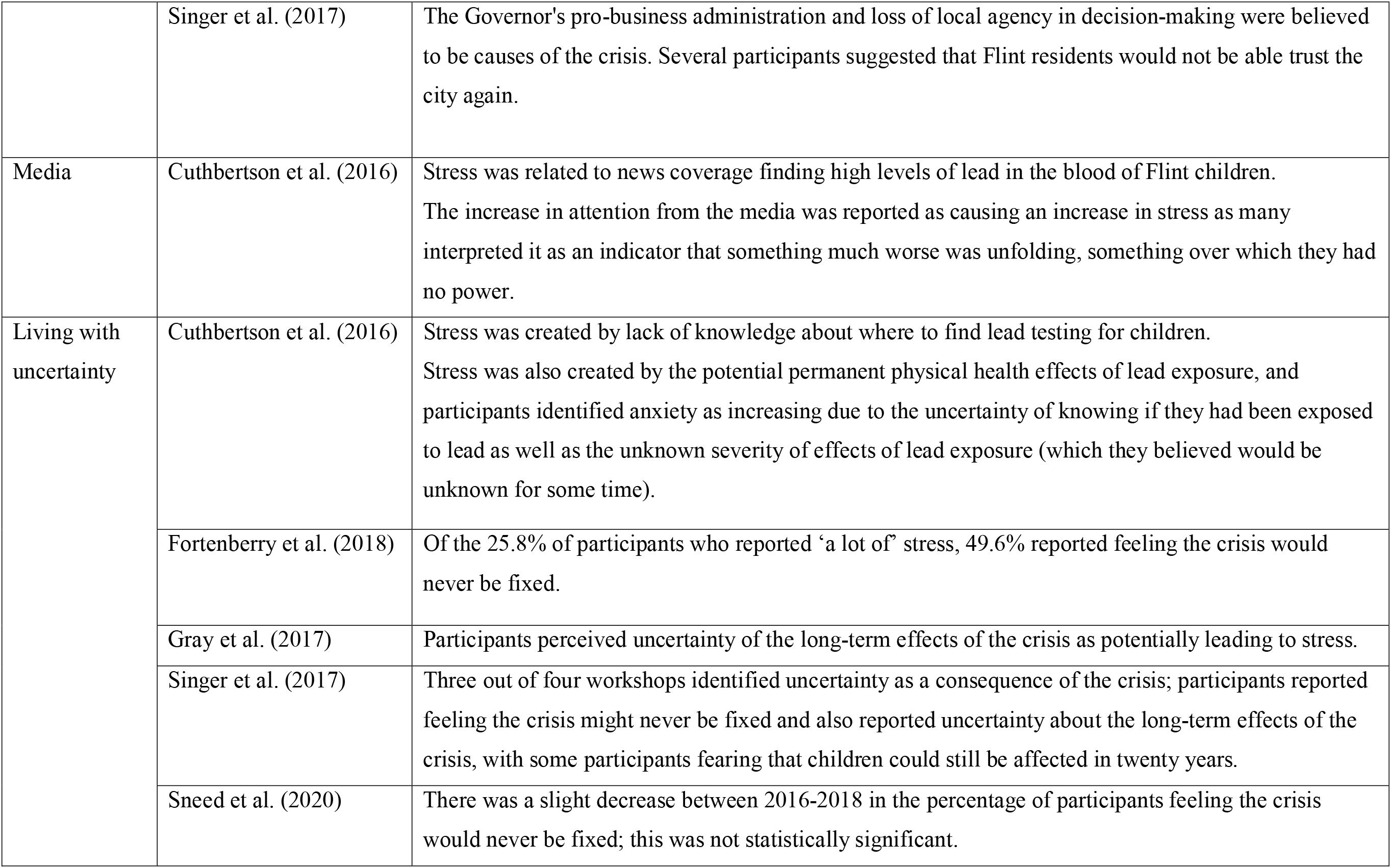
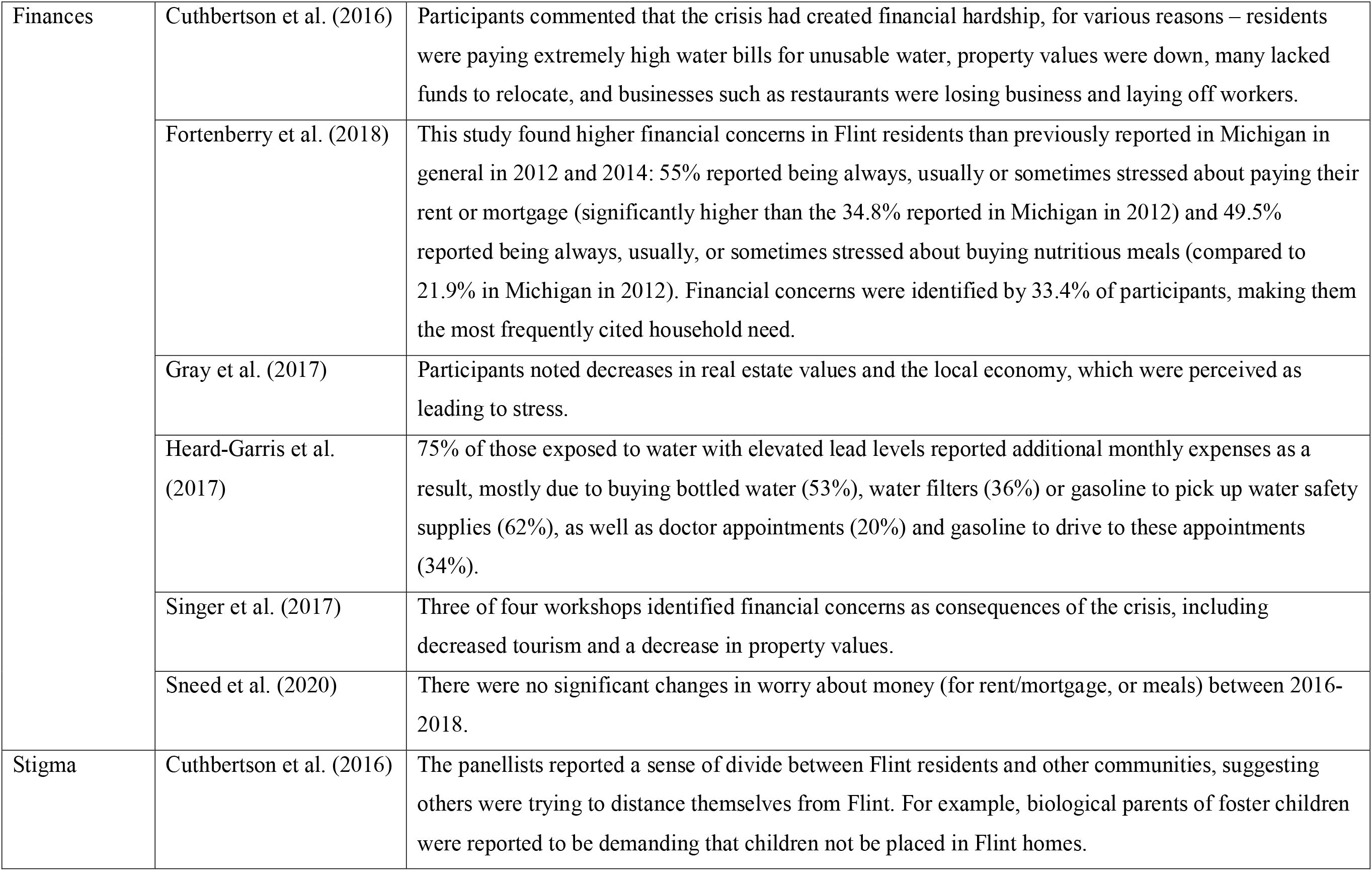
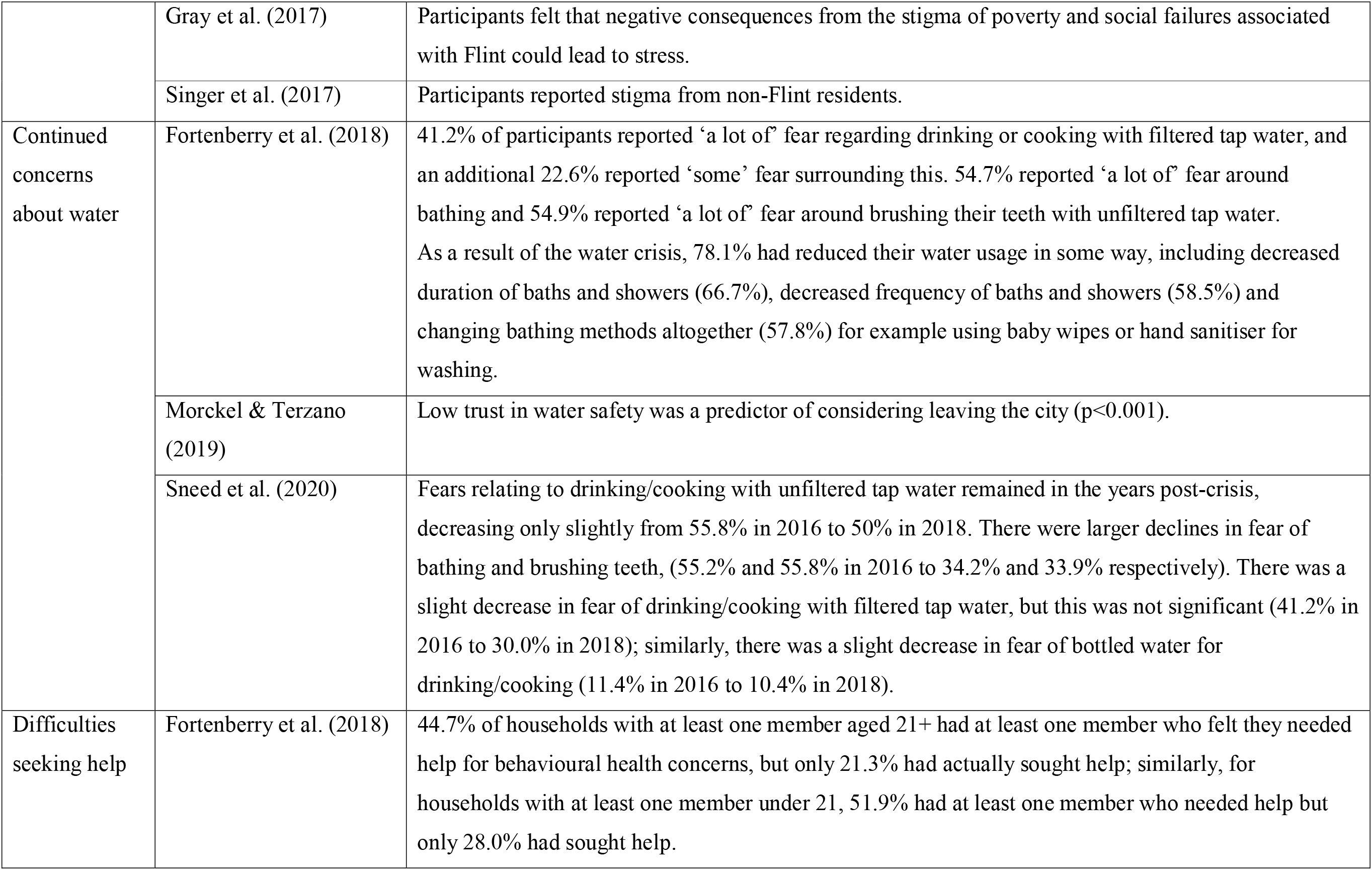
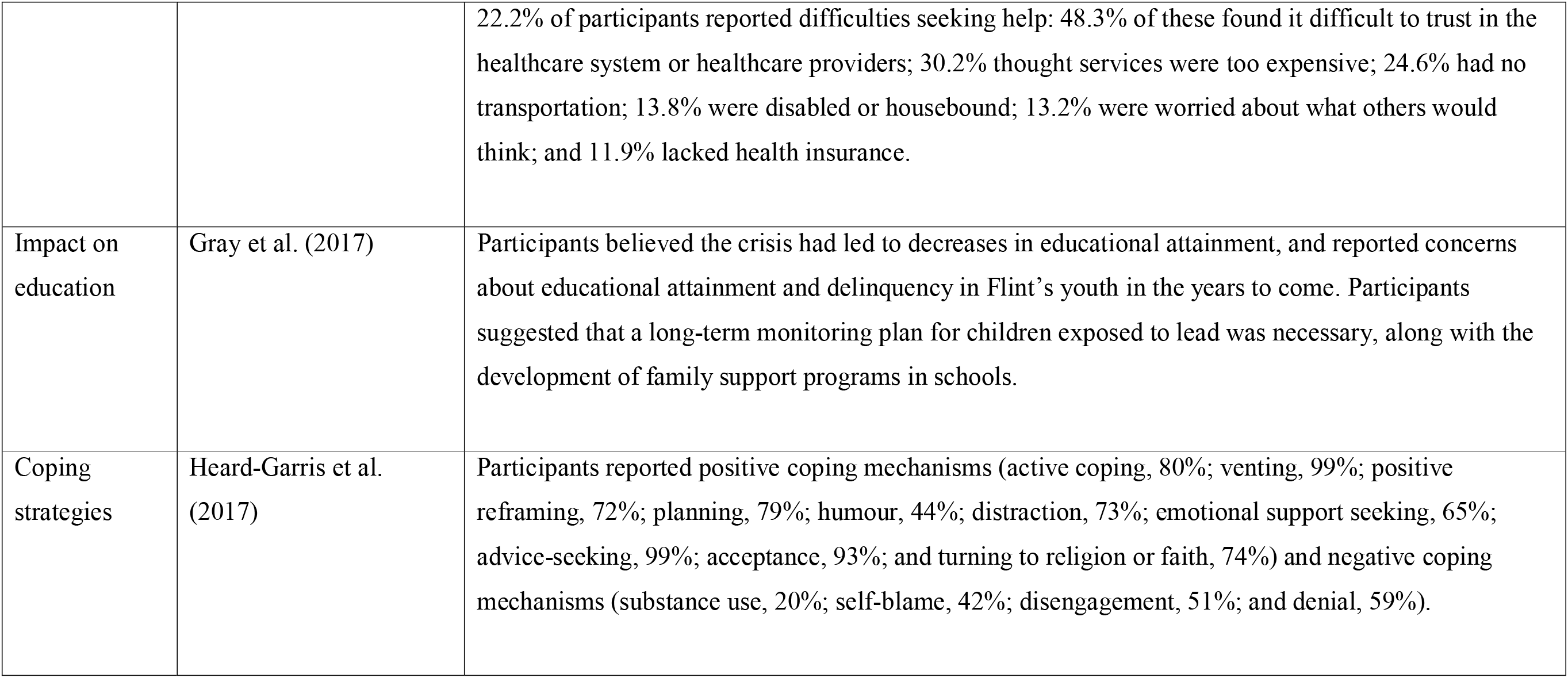
Themes emerging from included studies

### Impact on mental health

Participants directly affected by the water crisis reported symptoms of poor mental health in general (Kruger et al., 2017b), post-traumatic stress disorder (Kruger et al., 2017a), depression (Fortenberry et al., 2018), anxiety or stress (Fortenberry et al., 2018; Gray et al., 2017; Singer et al., 2017), sleep problems (Fortenberry et al., 2018; Kruger et al., 2018c), fear (Gray et al., 2017), aggressiveness (Fortenberry et al., 2018), trouble concentrating (Fortenberry et al., 2018), emotional outbursts (Fortenberry et al., 2018), decreased appetite (Fortenberry et al., 2018), and exacerbation of pre-existing health conditions (Gray et al., 2017). Cuthbertson et al.’s (2016) expert panellists believed the water crisis was increasing stress, anxiety and to a lesser extent depression among Flint’s population, and that residents had been left feeling angry, defeated and on edge. They also believed that effects of stress were not limited to Flint residents and that those outside of Flint could also be stressed due to knowing people affected by the crisis.

A study comparing indicators of psychological wellbeing in Flint after the water crisis with similar indicators in Michigan before the crisis (Fortenberry et al., 2018) found higher negative quality of life indicators in Flint after the crisis than in both 2012 and 2014, as well as significantly higher poor mental health and a significantly higher number of people reporting that physical or mental health had limited their usual activities than in 2013–2015.

More positively, Sneed et al.’s (2020) study showed some improvements in health outcomes over the years post-crisis: for adults aged 21 and over, aggressiveness, depressed mood, emotional outbursts, and anxiety/stress were lower in 2018 than they were in 2016, and for those aged under 21, there were significant declines in aggressiveness, emotional outbursts and problems in school. However, there were no significant changes in reports of trouble concentrating, decreased appetite or sleep problems for those aged 21 and over; or sleep problems, decreased appetite, depressed mood or anxiety/stress for those under 21.

### Risky behaviours

Kruger (2018) found that those who experienced poorer quality tap water demonstrated higher rates of risk-taking behaviour: in particular, poorer water quality experiences were associated with higher likelihood of tobacco smoking, higher self-reported HIV risk, higher likelihood of being involved in a physical fight and less healthy diet. Increased substance use was also noted by Heard-Gariss et al. (2017) and increased use of nicotine products by Fortenberry et al. (2018). Sneed et al. (2020) found no significant decrease in substance use over the years after the crisis, 2016–2018. Cuthbertson et al.’s (2016) participants believed that the mental health effects of the crisis could spill over into other areas of behavioural health, and could increase abuse, alcohol misuse, illicit drug use and prescription drug misuse.

### Demographic factors associated with mental health outcomes

Respondents in Cuthbertson et al.’s (2016) study felt that the entire community would be affected by the stress of the crisis, but particularly those in low-income African-American populations. Kruger et al. (2017a) found that younger age and fewer years of continuous education were associated with higher PTSD symptoms and likelihood of screening positive for PTSD.

### Distrust in public health officials

Cuthbertson et al.’s (2016) participants felt mental health consequences were related not only to the water contamination itself but to distrust of authority figures and lack of confidence in the government. Participants suggested the crisis had created distrust among city and state leadership; residents had lost trust in political officials and community leaders. This led to a sense of abandonment, due to no one taking responsibility for the water problems, which exacerbated mental health problems. Decreased trust in officials was also noted by Fortenberry et al. (2018), Gray et al. (2017), Kruger (2018), Morckel and Terzano (2019) and Singer et al. (2017). This decrease in trust may be due to feeling overlooked by decision-makers (Fortenberry et al., 2018). In terms of what could be done to restore trust in the government, Morckel and Terzano’s (2019) participants suggested removing local elected officials, more honesty from elected officials, improving transparency and communication with residents, and fixing the water situation. Worryingly, 7.9% of participants in this study did not know how trust could be restored, and 10.9% felt nothing could be done to restore it.

### Media

Stress was related to news coverage finding high levels of lead in the blood of Flint children (Cuthbertson et al., 2016). In the same study, the increase in attention from the media was reported as causing an increase in stress as many interpreted it as an indicator that something much worse was unfolding, something over which they had no power.

### Living with uncertainty

Participants cited various sources of uncertainty leading to stress: lack of knowledge about where to find lead testing for children in order to assess the potential impact of the crisis on their children (Cuthbertson et al., 2016); uncertainty of knowing if they had been exposed to lead as well as the unknown severity of effects of lead exposure (Cuthbertson et al., 2016); feeling that the crisis was not over and would never be fixed (Fortenberry et al., 2018; Singer et al., 2017); and uncertainty of the long-term effects of the crisis (Gray et al., 2017; Singer et al., 2017). Sneed et al. (2020) found a slight decrease between 2016–2018 in the percentage of participants feeling the crisis would never be fixed; however, this was not statistically significant.

### Finances

Participants frequently reported financial concerns and hardships (Cuthbertson et al., 2016; Fortenberry et al., 2018; Gray et al., 2017; Heard-Garris et al., 2017; Singer et al., 2017). Financial concerns appeared to be higher in Flint residents after the crisis than previously reported in Michigan in 2012 and 2014 (Fortenberry et al., 2018). These increased concerns were due to Flint residents paying extremely high water bills for unusable water (Cuthbertson et al., 2016); decreased property values (Cuthbertson et al., 2016; Gray et al., 2017; Singer et al., 2017); lacking funds to relocate (Cuthbertson et al., 2016); decreases in the local economy (Cuthbertson et al., 2016; Gray et al., 2017); decreases in tourism (Singer et al., 2017); and additional monthly expenses such as buying bottled water and water filters, doctor appointments, and buying gasoline to travel to appointments or to pick up water safety supplies (Heard-Garris et al., 2017). Sneed et al.’s (2020) study showed no significant change in financial concern in the years post-crisis, 2016–2018.

### Stigma

The panellists in Cuthbertson et al.’s (2016) study reported a sense of divide between Flint residents and other communities, suggesting others were trying to distance themselves from Flint. For example, biological parents of foster children were reported to be demanding that children not be placed in Flint homes. Similarly, Gray et al.’s (2017) participants felt that negative consequences from the stigma of poverty and social failures associated with Flint could lead to stress and Singer et al.’s (2017) participants also reported experiencing stigma from non-Flint residents.

### Continued concerns about water

In Fortenberry et al.’s (2018) study, over 60% of participants reported fear regarding drinking or cooking with filtered tap water, and over half reported ‘a lot of’ fear around bathing and brushing teeth with unfiltered tap water. As a result of the water crisis, more than three quarters had reduced their water usage in some way, including decreased duration and frequency of baths and showers and changing bathing methods altogether, for example using baby wipes or hand sanitiser for washing. Morckel and Terzano’s (2019) participants also reported low trust in water safety, which was a significant predictor of considering leaving the city (p< 0.001). Sneed et al. (2020) found that fears related to drinking and cooking with tap water did not significantly decrease between 2016–2018; however, there were much larger decreases in fears of bathing and brushing teeth.

### Difficulties seeking help

Fortenberry et al. (2018) found that only approximately half of those participants who felt they needed help for behavioural health concerns actually sought help. They reported barriers to seeking help such as finding it difficult to trust in the healthcare system or healthcare providers; finding services too expensive; having no transportation; being disabled or housebound; concerns about what others would think; and lack of health insurance.

### Impact on education

Gray et al.’s (2017) participants believed the crisis had led to decreases in educational attainment, and reported concerns about educational attainment and delinquency in Flint’s youth in the years to come. Participants suggested that a long-term monitoring plan for children exposed to lead was necessary, along with the development of family support programs in schools.

### Coping strategies

The majority of Heard-Garris et al.’s (2017) participants reported using positive coping mechanisms such as active coping, venting, positive reframing, planning, humour, distraction, emotional support seeking, advice-seeking, acceptance, and turning to religion or faith. Negative coping mechanisms such as denial and disengagement were also reported by more than half of the participants, and over 40% reported self-blame.

## Discussion

The negative effects caused by the water crisis on Flint’s residents have created a variety of issues in the affected population. Studies found in this review suggest various degrees of anxiety, depression, post-traumatic stress, sleep problems and worries about physical health existing in the affected population. Additionally, fear, negative coping strategies such as smoking and alcohol misuse, and risky health behaviours were prominent findings as response reactions from those affected by the crisis. These negative consequences were exacerbated by lowered trust in public health and government officials, heightened uncertainty about the long-term impacts of the crisis and the appropriate course of action to resolve emerging issues, and increased amount of financial hardships caused by the crisis. Self-perceived stigma from others in the community and at-large population along with difficulties seeking help were beliefs found in this review to prevent those affected to improve their respective situation. This review also found evidence that concerns and doubt about the tap water in Flint, Michigan continued even after the state of emergency was lifted.

However, there are some positive implications from this review, as the major stressors identified can be targeted with interventions and consequently their impact lessened. For example, negative coping strategies such as smoking and alcohol misuse and risky health behaviours can be addressed in the Flint community through public health interventions and programs. Programs that enable peer and group support have been effective in establishing and enabling changes in these behaviors (Ford et al., 2013; Tracy & Wallace, 2016). Additionally, in a longitudinal study of the first three years after the crisis, improvements in mental health outcomes as well as a decline in fears of using tap water for bathing and brushing teeth were seen, which suggest public health messages surrounding water usage for these purposes were well-received (Sneed et al., 2020). However, other concerns related to the crisis remain unchanged, such as financial worries and an overall concern that the crisis will never be fixed. Use of evidence-informed interventions can reduce these concerns, enhance wellbeing, and improve functioning for affected individuals (Morganstein & Ursano, 2020). Despite progression in some beliefs, community efforts to reduce psychological distress are still warranted.

The psychological consequences of the Flint water crisis may generalise to other disasters, and support findings from similar incidents involving environmental contamination based on past disasters. For example, studies that examining the long-term mental health consequences of the Chernobyl nuclear power plant disaster show the affected populations developed an exaggerated sense of presumed exposure and danger due to Chernobyl over time, which fuelled an increased level of anxiety and perpetual stigmatization through the generations (Bromet et al., 2011; Havenaar et al., 2016; Samet & Patel, 2011, 2013). Besides similar results found in this review with impact on mental health and stigmatization, the themes of distrust in government officials and risky behaviours were also seen among populations affected by Chernobyl (Bolt et al., 2018; Samet & Patel, 2013). There were more depression, anxiety, and overall concerns specifically about children’s well-being among the affected populations even decades after the incident – this brings heightened urgency for further investigations and longitudinal studies related to the Flint Water Crisis to ensure something similar does not happen in Flint.

Additionally, the COVID-19 pandemic creates a possible compound effect to those affected by the Flint Water Crisis, as they now face coping with another emergency while still recovering from another. Despite being over three years since the incident, the communities, as seen in this review, are still reeling from the mental health impact which will be exposed further with the pandemic. Although coronavirus infects people regardless of income, the low-income communities in Michigan have been deeply affected with a higher number of cases than high-income ones, especially in Genesee County (Genesee County Health Department, 2020). This highlights the issue of how Michigan is segregated by income and race, with Flint a good representative of this in the state (Muhammad et al., 2018; Pulido, 2016), with race a crucial factor in the high caseload of confirmed COVID-19 patients in Flint (Fonger, 2020).

There are several implications for policy and practice based on the findings of this review. First, rebuilding trust in official communication and science within the Flint community will enhance ongoing efforts to resolve the widening gaps that were caused by the initial crisis. Local government and public health officials need to ensure they regain the trust of residents along with engaging and involving the Flint community and its members in recovery efforts. For example, ensuring transparency, seeking community input, and enabling two-way communication with the public on resolving issues will allow trust to be built institutionally. Having government and public health officials being held accountable, demonstrating integrity by admitting to mistakes, and seeking input from respected outside experts will bridge their past leadership woes and connect with the communities they are serving (Boufides et al., 2019).

In Flint, efforts are being made to provide urgent mental health services. Psychological first aid training for people interested in helping others cope with the water emergency has been provided by the Flint Community Resilience Group (Cuthbertson et al., 2016) and the Flint RECAST program educates residents about trauma (Ahmad, 2019). However, there remain concerns about whether enough is being done and also whether people would actually seek psychological help (Goodnough & Atkinson, 2016). Still, mental health services need to be readily available and ramped up in the Flint community. Government and public health officials should identify and strengthen resources for mental health services for affected residents, and conduct follow-up mental health assessments to evaluate change over time. Additional mental health interventions should also occur in Flint, especially to include the current COVID-19 pandemic. However, economic factors, such as access and costs, need to be considered when implementing mental health interventions. An increase of educational media campaigns and handouts can also enhance the mental health interventions by emphasising the potential long-term mental health implications of the crisis, providing contact information for support groups and readily available mental health services. All trust-building and mental health services done in the Flint community will build infrastructure and enhance the efforts if future crises arise.

Future research in Flint should go beyond the profound societal effects caused by the crisis and create opportunities to resolve the disadvantages for the Flint community to become an example of a community that can bounce back and be resilient from future public health emergencies. Despite the concept of community resilience having been variously defined by researchers, government officials and public health practitioners without a unifying meaning, several themes found across the definitions align with the results found in this review and help to prioritize future research (Patel et al., 2017). Community participatory research examining crisis communication, leadership and resources can help build trust and evidence-based infrastructure and working relationships between officials and Flint community members. With a compound effect of the current impact caused by the COVID-19 pandemic, maladaptive coping strategies and risky health behaviours may need urgent attention to manage the distressing emotions and community memory of compounded crises (Morganstein & Ursano, 2020). Facing multiple public health emergencies can cause inherent challenges and trust for communities but amplifying the evidence-based interventions and mental health research can not only address the current public health emergencies but can also help prevent a bigger distress caused by future ones as well.

### Limitations

The majority of studies reviewed were cross-sectional and thus suggest associations, rather than causality; very few studies were longitudinal, so the long-term effects of the Flint Water Crisis are unclear. Much of the included research was conducted while the crisis was ongoing: the full impact of the crisis may therefore be under-estimated, due to the speed with which data was collected; conversely, the mental health consequences could be overestimated as Sneed et al.’s (2020) findings suggest that symptoms may improve over time. The majority of studies did not compare rates of mental health problems between Flint and other populations, or pre-Water Crisis rates in Flint, making it difficult to truly ascertain the impact of the crisis on mental health. Other factors, such as the economic recession or personal circumstances, could also have affected mental health. Additionally, much of the data in the included papers was obtained via self-reports, which may not necessarily be reliable. In terms of the review process itself, the main limitation is that screening and data extraction was done by only one author: the review could be improved by a second author carrying out the same process to ensure consistency. Secondly, the decision to include only peer-reviewed literature and not grey literature may mean that the data reviewed in this article is subject to publication bias.

### Conclusion

Literature on the impact of the Flint Water Crisis suggests considerable psychological consequences to Flint residents, exacerbated by mistrust in officials and financial difficulties. While Flint struggles to recover from this crisis, the city has also seen a much higher rate of COVID-19 than other parts of Genesee County, meaning Flint residents now have two disasters to cope with. Our review highlights the urgent need for more mental health resources for the people of Flint.

## Data Availability

All data is present in the manuscript.

